# Protocol For a Scoping Review of Patient-Preferred Outcomes in Depressive Disorders

**DOI:** 10.1101/2024.12.20.24319426

**Authors:** Megan A Boreham, Mariia Bogdanova, Madeleine Bloomfield, Anthony J Cleare, Rebecca Strawbridge

## Abstract

Depressive disorders remain a leading cause of health-related burden globally. Patients often suffer with symptoms throughout their lifetime, and rates of remission have been estimated to be as high as 66%. Clinicians typically favour the use of scales monitoring quantity and severity of depressive symptoms to evaluate response to treatment and determine whether patients have entered remission. However, given the complexity of the disorders and the subjective nature of mental health, research indicates that patients value outcomes beyond this narrow definition, identifying outcomes related to functioning and quality of life as important in addition to control of depressive symptoms. These studies are typically limited by relatively small sample sizes and associated lack of generalisability.

Here, we outline the protocol for a scoping review that aims to comprehensively collect and review research into patient preferred outcomes in treatment and management of depressive disorders. Our review was designed in accordance with the PRISMA Extension for Scoping Reviews (PRISMA-ScR) statement. Embase, Medline, PsycINFO, and CINAHL will be searched for relevant records. Qualitative and mixed-methods studies will be included, and all data will be synthesised and presented in narrative form. We anticipate that our results will provide useful insight for clinicians when caring for patients with depressive disorders and highlight potential misalignment between patient and clinician treatment goals.

## BACKGROUND

Mental disorders, of which depressive disorders are considered the most disabling, are within the top 10 leading cause of health-related burden across the globe^1^. Burden for patients remains high across the lifetime, and outcomes with current treatment interventions for major depressive disorder remain suboptimal: the STAR*D trial found approximately only two thirds of patients achieved remission after 1 year of tailored pharmacological or psychological treatment, suggesting that one third of patients can be classified as treatment resistant^2^. The most clinically utilised treatment goals for depressive disorders are often centred on symptomatic control, denoted by changes in the quantity and severity of depressive symptoms measured using either patient or clinician-administered rating scales. However, using the above principle to define remission from depressive disorders may not be the optimal approach when considering the following two factors: firstly, depressive disorders have high rates of recurrence^3^, rendering full remission an unrealistic goal in such patients who are likely to experience residual deficits in other domains (e.g., social functioning); secondly, whilst helpful for objective measurement of outcomes in randomised control trials, these outcome measures do not inherently reflect patient views and values pertaining to optimal management of their disorder^4^.

The need to identify outcomes relevant and important to patients is exemplified in healthcare systems such as the National Health Service (NHS), where a shift from evidence-based practice towards value-based practice in commissioning of mental health approaches is central to the NHS 10 Year Plan^5^. Value-based commissioning places a strong focus on consideration of outcomes that matter to service users, and involvement of users in coproduction of healthcare services^6^. Recent evidence has started to suggest that the tools currently used in the diagnosis and management of depressive disorders, typically developed over 25 years ago, fail to adequately capture measures and outcomes that are of importance to patients. The little research into patient-preferred outcomes indicates that patients value a broad range of indicators beyond symptomatic control in monitoring of their depression, including a return to a normal level of functioning (both social and occupational), optimism, and deriving enjoyment from activities^7-10^. Discrepancies in views between clinicians and patients can lead to differences in treatment expectations, which may lead to disappointment and dissatisfaction from the patient.

Here, we outline the protocol for a scoping review that aims to comprehensively summarise research into outcomes and measures identified as important to patients with depressive disorders. We anticipate that concepts related to functioning and a ‘return to everyday life’ will be identified as important to patients, although it is unclear if this will differ when patients consider longer-term vs short-term goals. The findings of this review will thus provide useful insight for clinicians when considering treatment strategies for patients, particularly in evaluation of benefit of treatment strategies.

## OBJECTIVE & AIMS

To review and summarise existing research into patient-preferred outcomes and treatment goals in treatment and management of depressive disorders. Given the limited size of the literature to date, we plan a broad and inclusive review not limited by specific interventions or outcomes focused on. The specific questions are as follows, although we appreciate that the review findings are largely dependent on the available evidence and thus not restricted to these questions.

- Primary research questions:
  - How do patients describe or define remission, recovery or adequate management of a depressive episode?
  - What do patients characterise as meaningful outcomes or treatment goals for a depressive episode?
- Ancillary aims:
  - How do these definitions and descriptions vary across different depressive disorders?
  - Are there variations in definitions or descriptions when patients consider either pharmacotherapy or psychotherapy?
  - Do these definitions or descriptions change when patients consider the short vs long term outcomes?
  - Are there variations in the definitions or descriptions between patients currently experiencing a depressive episode and those providing retrospective perspectives?
  - Do patients prefer their own descriptions or definitions, or those provided by clinicians?
  - Do these patient perspectives translate into specific patient-preferred clinical instruments, with a preference for either patient-reported or clinician-reported?

## SEARCH STRATEGY AND INFORMATION

A scoping approach was selected in lieu of a systematic review design due to the breadth of our research questions and our aim to summarise all the available literature. We will search several sources for published and unpublished records using relevant keywords and databases: Embase, Medline, PsycINFO, and CINAHL. No time restrictions will be applied; language will be restricted to English due to lack of resources for translation. Reference lists of resulting records (including studies and related reviews) and reference lists from notable authors will be inspected by hand to retrieve any additional relevant records.

## Search strategy for Medline (OVID)

MeSH terms (all exploded): (depression OR mood disorder) AND patient preference AND (treatment outcome OR quality of life OR patient reported outcome measures OR mental health recovery) AND (qualitative research OR interview OR focus groups OR surveys and questionnaires)

Free text search [all fields]: (depression OR depressive* OR depressed OR MDD or TRD).mp AND ((relevant OR important OR preferred OR perspective* OR preference* OR view* OR meaningful OR choice OR choose OR chosen OR chose OR favouri* or favori* OR favoured OR favored OR affinity OR desire* OR select*).mp AND adj5 (outcome* or measure* OR endpoint* OR parameter* OR recovery OR remission OR improvement* OR treatment goal* OR therap* goal*).mp AND adj5 (patient* OR service user*).mp)) AND (questionnaire* OR survey* OR interview* OR focus group* OR conversational analys* OR qualitative OR mixed method*).mp

The same strategy will be used across all other databases after syntax adaptation.

## STUDY SELECTION

All qualitative and mixed methods studies investigating the patient perspective on definitions or descriptions of recovery or remission from, or adequate management of depressive episodes will be included. Patients will be restricted to adults over the age of 18 with clinically significant symptoms of depression and include either those experiencing a current episode or those reporting retrospectively on prior episodes. Any qualitative or mixed-methods study design will be included. No restrictions will be placed over the intervention patients’ may currently be receiving, the healthcare setting, or the geographical location. Articles that describe caregiver or healthcare professional perspectives alone and not in combination with patient perspectives will be excluded. Conference abstracts will be included only if there is adequate data available; editorials and commentaries will be excluded.

Duplicate records retrieved will initially be de-duplicated using Ovid’s built-in tool for searches in Medline, Embase, and PsycINFO, and subsequently in Rayyan open-source review management software^11^. All records will be imported into Rayyan, to record the status of each article during the screening and inclusion process. Two authors (M.A.B & either M.Bl or M.Bo) will independently perform the abstract screening of the titles retrieved by the search. Articles included by at least one author will be eligible for full-text screening. Full-text screening will be carried out independently by two authors. Disagreement will be resolved by consensus or by consultation with a senior member of the review team (R.S).

## DATA SELECTION AND CHARTING

Two investigators will systematically extract data from each included record using a standardised data extraction tool designed for this review (Excel software). Data extracted will include bibliography (e.g., authorship, publication), methodological characteristics (study design, location, setting, research objectives or aims, participant population) and results (key findings and conclusions). Differences will be resolved as outlined above. Should further information or data be deemed necessary, an attempt to contact the original authors will be made. Although critical appraisal of evidence sources is not formally required in scoping reviews^12^, we aim to critically evaluate the studies for methodological quality and risk of bias where possible. We plan to use the following checklists: JBI Checklist for Qualitative Research^13^, JBI Checklist for Systematic Reviews and Research Synthesis^14^, and the Mixed Method Appraisal Tool^15^. The authors appreciate the domains evaluated will depend on the specific study design of the included records.

## DATA SUMMARY AND SYNTHESIS OF RESULTS

In accordance with the review aims and questions outlined above, we are interested in any description or definitions of recovery, remission, and treatment goals as provided by patients, to identify outcomes relevant to patients with depressive disorders. Methodological/bibliographic characteristics and key findings/results will be summarised using tables and text. Results will be compiled using a narrative synthesis, organised via the specified review questions outlined above. The narrative synthesis will explore patterns/themes arising from the included records, incorporating trend observations identified from the above outcomes where possible. The screening process will be documented in a PRISMA flow diagram to facilitate reproducibility, with reasons for exclusion being detailed.

## PROTOCOL AND REGISTRATION

This protocol was drafted in accordance with the PRISMA Extension for Scoping Reviews (PRISMA-ScR) statement^16^ and the JBI Manual for Evidence Synthesis^17^. The protocol will be registered and made available online via the medRxiv.org database.

## DATA AVAILABILITY

All data produced in this study will be available upon reasonable request to the authors.

